# Direct Oral Anticoagulants for Stroke Prevention in Atrial Fibrillation: A Global Synthesis of Randomized Evidence through Network Meta-Analysis

**DOI:** 10.1101/2025.05.18.25327653

**Authors:** Manyata Srivastava, Annu Gulia, Kamalesh Kumar Patel, Ashish Datt Upadhyay, Deepti Vibha, Pradeep Kumar

## Abstract

**Background and Objectives:** Direct oral anticoagulants (DOACs) have become preferred alternatives to vitamin K antagonists (VKAs) for stroke prevention in both valvular (VAF) and non-valvular atrial fibrillation (NVAF). However, comparative evidence on the relative efficacy and safety of different DOACs remains limited. This network meta-analysis aimed to systematically assess and compare the efficacy and safety of various DOACs versus VKAs in preventing stroke and related outcomes among patients with AF.

**Methods:** A comprehensive literature search of PubMed, EMbase, and the Cochrane Library was conducted for randomized controlled trials (RCTs) published up to October 31, 2024. A Bayesian network meta-analysis was performed using odds ratios (ORs) with corresponding 95% credible intervals (CrIs).

**Results:** A total of 43 RCTs were included, comprising 30 on VAF and 13 on NVAF. Compared to VKA, apixaban (OR = 0.81; 95% CrI: 0.73 to 0.91), dabigatran (OR = 0.77; 95% CrI: 0.68 to 0.87), and rivaroxaban (OR = 0.87; 95% CrI: 0.79 to 0.96) were significantly associated with reduced risk of IS/SE, while edoxaban showed a non-significant effect. All DOACs were significantly superior to VKA in reducing the risk of HS. For major bleeding, apixaban (OR = 0.69; 95% CrI: 0.55 to 0.88) showed a significant advantage over VKA, while dabigatran and rivaroxaban were associated with non-significant increases. In terms of all-cause mortality, apixaban significantly reduced risk in NVAF (OR = 0.88; 95% CrI: 0.82 to 0.96), whereas dabigatran, edoxaban, and rivaroxaban showed non-significant associations. In VAF, neither dabigatran nor rivaroxaban demonstrated a significant impact on mortality.

**Conclusion:** This network meta-analysis suggests that apixaban and dabigatran are associated with significantly better outcomes in terms of reducing ischemic and hemorrhagic events, major bleeding, and mortality compared to VKAs, particularly in NVAF patients. Edoxaban and rivaroxaban showed favourable but generally non-significant associations. These findings support the use of apixaban and dabigatran for stroke prevention in patients with AF.

## Introduction

Atrial fibrillation (AF) is a prevalent cardiac arrhythmia affecting over 33 million people worldwide and is a major contributor to cardioembolic stroke.^1–4^ Patients with AF carry a nearly fivefold increased risk of ischemic stroke, making long-term anticoagulation essential for secondary and primary stroke prevention.^4^ Traditionally, vitamin K antagonists (VKAs) such as warfarin have been the mainstay of treatment, but their use is challenged by inter-individual variability, frequent INR monitoring, dietary restrictions, and drug interactions.^5^

The emergence of direct oral anticoagulants (DOACs)—including apixaban, dabigatran, rivaroxaban, and edoxaban—has transformed anticoagulation strategies in AF. DOACs offer fixed dosing, rapid onset of action, and a more favourable safety profile, especially with respect to intracranial hemorrhage.^5,6^ Randomized controlled trials (RCTs) such as RE-LY,^7^ ROCKET-AF^5^, ARISTOTLE,^6^ and ENGAGE AF-TIMI 48^8^ have established the non-inferiority or superiority of individual DOACs compared to VKAs for preventing stroke and systemic embolism (SE) in non-valvular AF (NVAF). However, most of these trials excluded patients with valvular AF (VAF), leaving a gap in evidence for this subpopulation.

While several pairwise meta-analyses have compared DOACs with VKAs, they are limited in scope and unable to provide a comprehensive comparative ranking across all available agents.^9^ Moreover, prior analyses have often combined heterogeneous populations, lacked stratification by AF subtype (VAF vs. NVAF), or focused on a single outcome, failing to integrate data across multiple clinically relevant endpoints such as ischemic stroke/systemic embolism (IS/SE), hemorrhagic stroke (HS), major bleeding, and all-cause mortality^10–12^.

Given these limitations, a robust synthesis of both direct and indirect evidence is necessary to guide clinical decision-making. Network meta-analysis (NMA)^13^ offers a powerful approach to compare multiple interventions simultaneously, allowing the integration of RCT data and the generation of a relative ranking of treatments. This study aims to conduct a comprehensive NMA of RCTs to evaluate the comparative efficacy and safety of DOACs versus VKAs for stroke prevention in patients with AF, stratified by VAF and NVAF. By addressing the heterogeneity and gaps in existing literature, this analysis seeks to inform evidence-based prescribing practices and optimize anticoagulant selection in diverse AF populations.

## Methods

### Registration

This systematic review and network meta-analysis (NMA) adhered to the 2020 guidelines outlined in the Preferred Reporting Items for Systematic Reviews and Meta-Analyses-Network meta-analysis (PRISMA-NMA).^14^ Our protocol for this network meta-analysis was registered on PROSPERO under the registration number **CRD42023495164**.

### Inclusion Criteria

The PICOS framework was used to establish eligibility for inclusion

**(P)– Population**: Stroke patients diagnosed with atrial fibrillation (AF).

**(I)– Intervention**: Subjects randomized to DOACsor VKAs groups once, twice times per day.

**(C)– Comparator**: Any another DOACs or VKAs group

**(O)– Outcome**:Ischemic stroke (IS) or systemic embolism (SE), hemorrhagic stroke (HS), major bleeding and all-cause mortality

**(S)– Study Design:** Randomized controlled trials (RCTs)

### Exclusion criteria

1. Review articles, letters, dissertations report, editorials, conference abstracts, case reports, case series, and other non-original research publications.
2. Studies not reporting outcomes related to the risk of stroke.
3. Studies that conducted on animal subjects.
4. Duplicate publications.

### Search Strategy

A comprehensive literature search was conducted across major medical databases, including PubMed, EMBASE, and the Cochrane Library, covering publications up to October 31, 2024. To ensure thorough coverage, reference lists of all relevant articles and previous meta-analyses were also manually screened. The search strategy was designed to identify studies evaluating the effectiveness of direct oral anticoagulants (DOACs) for stroke prevention. Keywords and MeSH terms included: *“oral anticoagulants” OR “DOACs” AND “stroke prevention” OR “ischemic stroke” OR “systemic embolism” OR “hemorrhagic stroke” AND “atrial fibrillation” OR “non-valvular atrial fibrillation” AND “VKAs” AND “major bleeding” OR “all-cause mortality” OR “systemic embolism”*. Detailed search strategies are provided in **Supplementary Table S1**.

### Study Selection and Data Extraction

Two reviewers (MS and AG) independently screened the titles and abstracts of identified records to determine eligibility. Full-text articles of potentially relevant studies were then assessed against the inclusion criteria. Data were extracted using a standardized form, capturing: first author, publication year, study location, participant ethnicity, study design, duration, sample size, treatment regimens, drug dosage and frequency, follow-up duration, and clinical outcomes. Disagreements were resolved through discussion, with arbitration by the corresponding author (PK) if needed.

### Quality Assessment

The methodological quality of included RCTs was evaluated using the modified Jadad scale.^15,16^ This tool assesses eight domains: randomization, adequacy of randomization, blinding, adequacy of blinding, handling of withdrawals/dropouts, inclusion/exclusion criteria, reporting of adverse effects, and appropriateness of statistical analysis. Scores range from 0 to 8, with higher scores indicating better quality.

### Statistical Analysis

Using a Bayesian random-effects model, we performed a pairwise meta-analysis of direct comparisons in order to obtain direct evidence. Odds ratios (OR) and 95% credible intervals (CrI) for dichotomous outcomes were used to estimate treatment effects. Based on the observed data, credible intervals indicate a range of feasible OR values. The Surface under the Cumulative Ranking (SUCRA) curve values, along with their CrI, were calculated as well in order to evaluate the uncertainty in the DOACs or VKAs rankings. A SUCRA value closer to 100% indicates a higher rank in DOACs compared to VKAs in the NMA. Online web based MetaInsight tool (MetaInsight V4.0.2 Beta software) for Bayesian network meta-analyses were employed using random-effects model for all comparisons.^17^

## Results

### Study Selection

The initial database search identified 860 records. Following the removal of 183 duplicates, a total of 558 unique records were screened based on titles and abstracts. Of these, 97 full-text articles were assessed for eligibility. After applying predefined inclusion and exclusion criteria, 43 studies were deemed suitable and included in the final systematic review and network meta-analysis.

The study selection process is detailed in **Figure 1**, which presents the PRISMA-NMA flow diagram, illustrating each stage of screening and inclusion.

**Figure 1:**
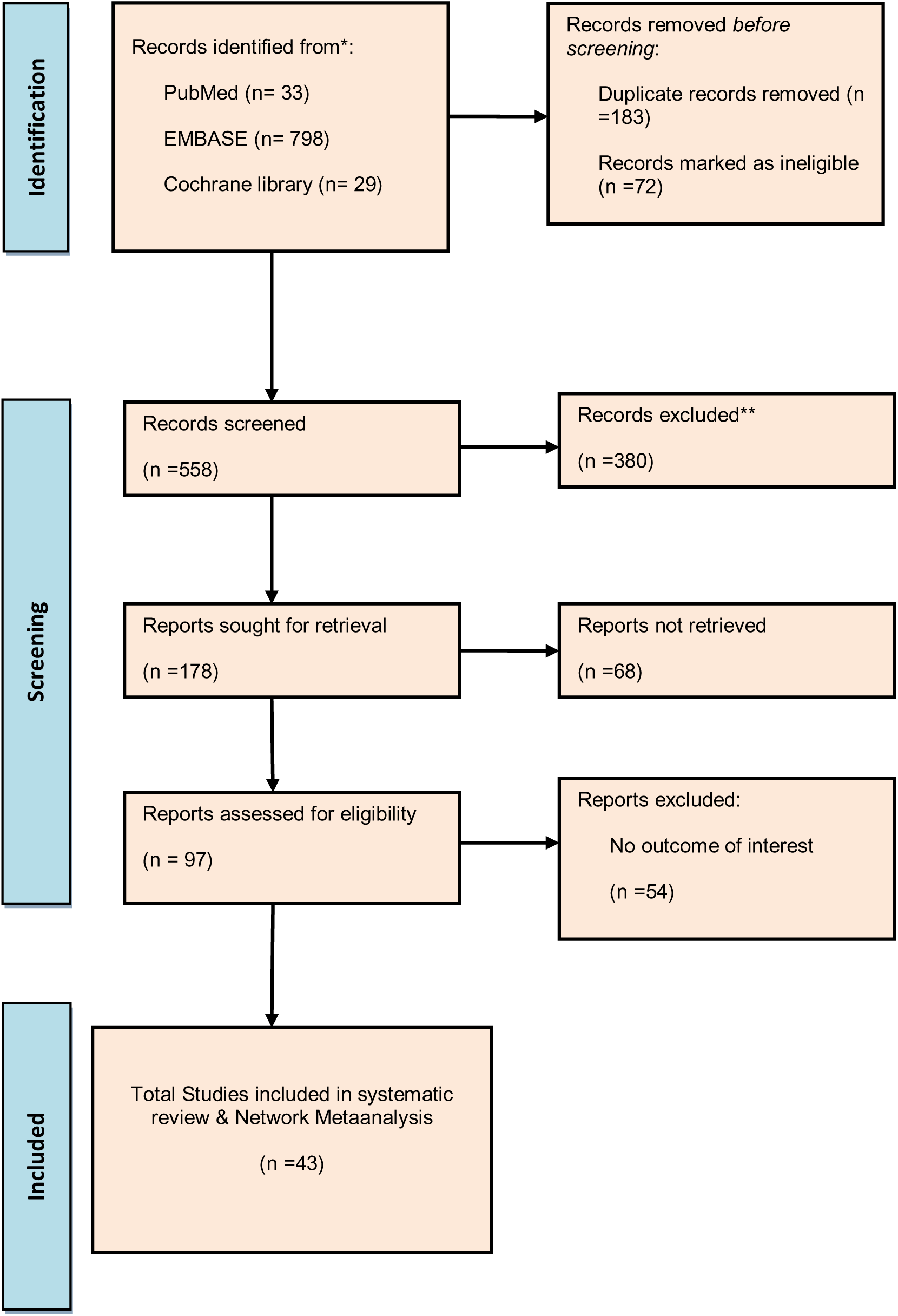
PRISMA Flow diagram for the selection of studies and specific reasons for exclusion from the present network meta-analysis

### Characteristics of Included Studies

A total of 43 RCTs^6–8,13–53^ were included in the network meta-analysis, encompassing a broad range of clinical settings, populations, and outcomes. These studies evaluated the efficacy and safety of four DOACs—dabigatran, rivaroxaban, apixaban, and edoxaban—in comparison to VKAs or placebo. The publication years of the included studies spanned from 2007 to 2022, reflecting an extensive temporal scope and covering both early and more contemporary trials in anticoagulation therapy.

Among the 43 included RCTs,^6–8,13–53^ 31 reported outcomes related to IS, 20 reported on HS, 38 assessed major bleeding, and 27 provided data on all-cause mortality. These endpoints allowed for a comprehensive comparison of the benefit-risk profiles of DOACs relative to VKAs and placebo.

To enable a full synthesis of both direct and indirect comparisons across the five intervention groups (four DOACs and one control group), we employed a frequentist network meta-analysis. This approach facilitated the estimation of relative treatment effects and enabled ranking across outcomes based on the available evidence from the included trials.

Regarding the study populations, 13 RCTs included patients with NVAF, while 30 enrolled patients with VAF. This variation in AF type reflects the heterogeneity of the clinical populations assessed and is important in interpreting treatment effects, as the thromboembolic risk and response to anticoagulants can differ between these subgroups.

The network plots presented in Figure 2 (a–d) illustrate the structure and connectivity of the available evidence for each clinical outcome: (a) IS/SE, (b) HS, (c) major bleeding, and (d) all-cause mortality. Each outcome is analyzed across three population groups: VAF, NVAF, and the combined (NVAF + VAF) population. Across all networks, most treatment comparisons were centered on DOACs (dabigatran, apixaban, rivaroxaban, edoxaban) versus VKAs, with fewer direct comparisons among DOACs themselves. The size of the nodes reflects the number of patients receiving each treatment, while the line thickness denotes the number of trials contributing to each direct comparison. The networks were well-connected for IS/SE and major bleeding, while fewer studies directly informed HS and mortality outcomes in the VAF subgroup. The observed network structure supported robust direct and indirect comparisons for the ensuing NMA.

**Figure 2.**
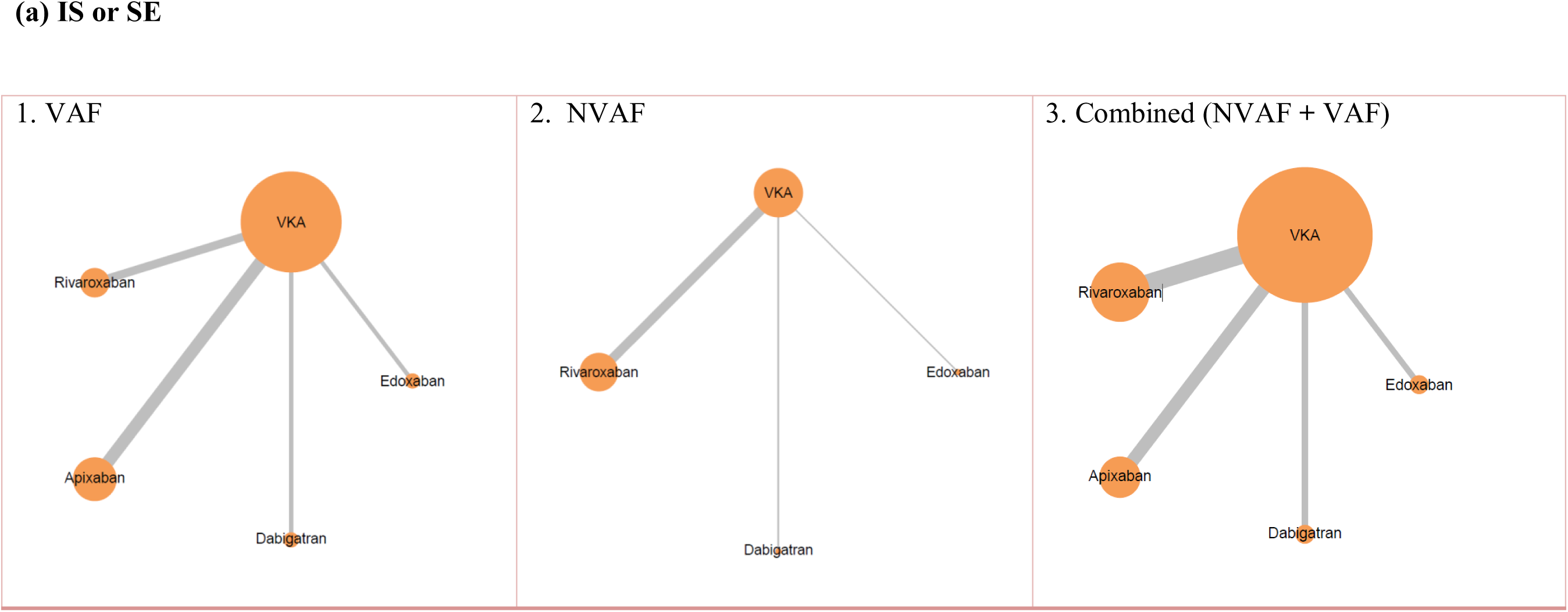
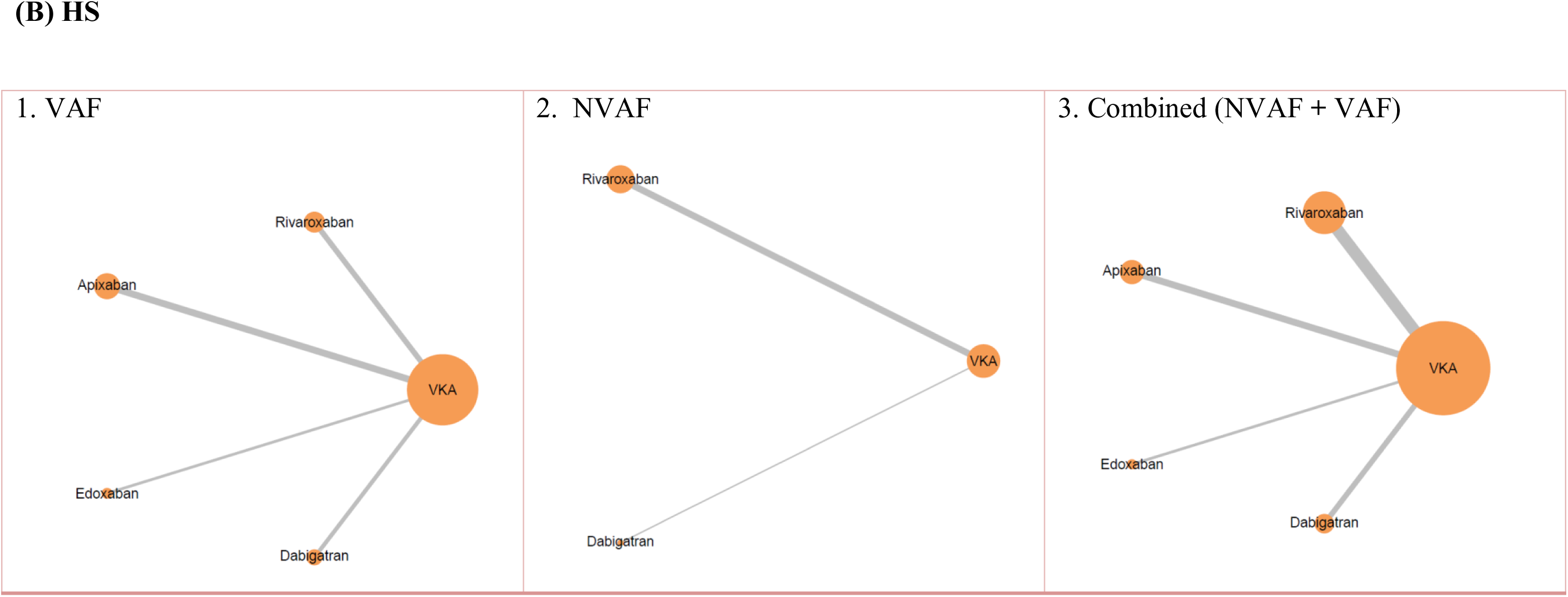
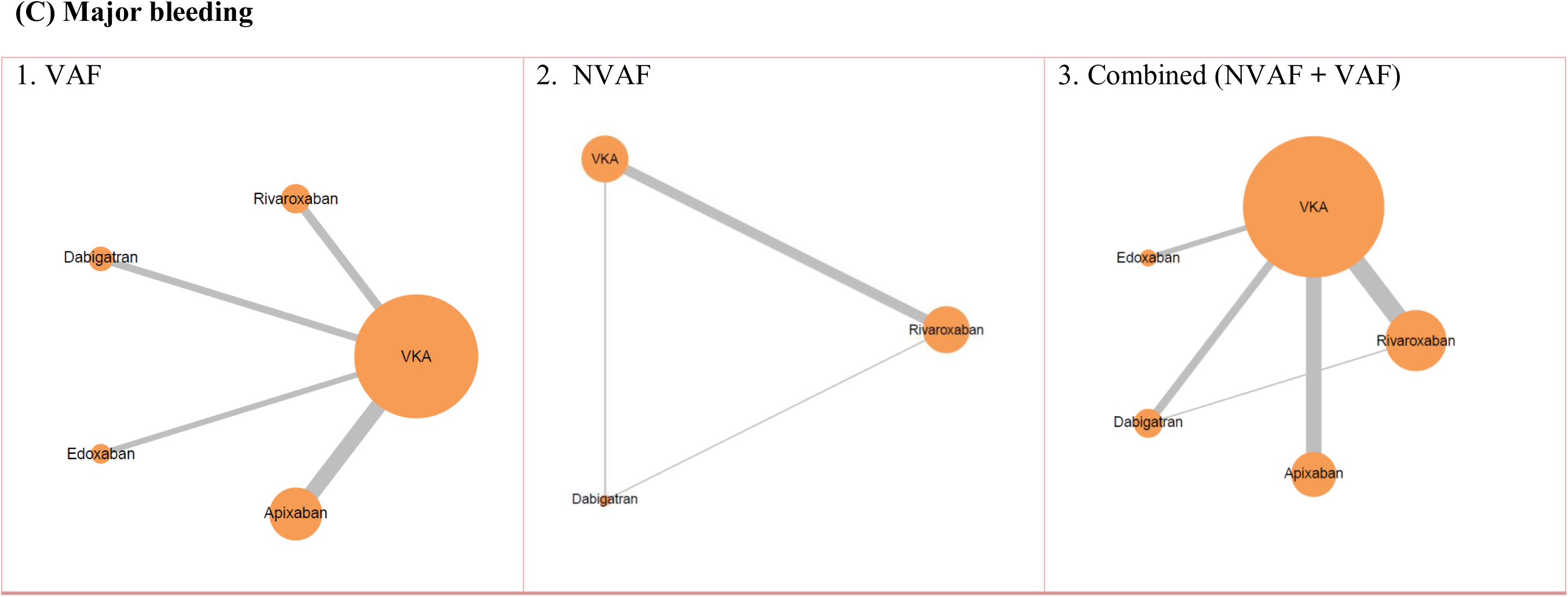
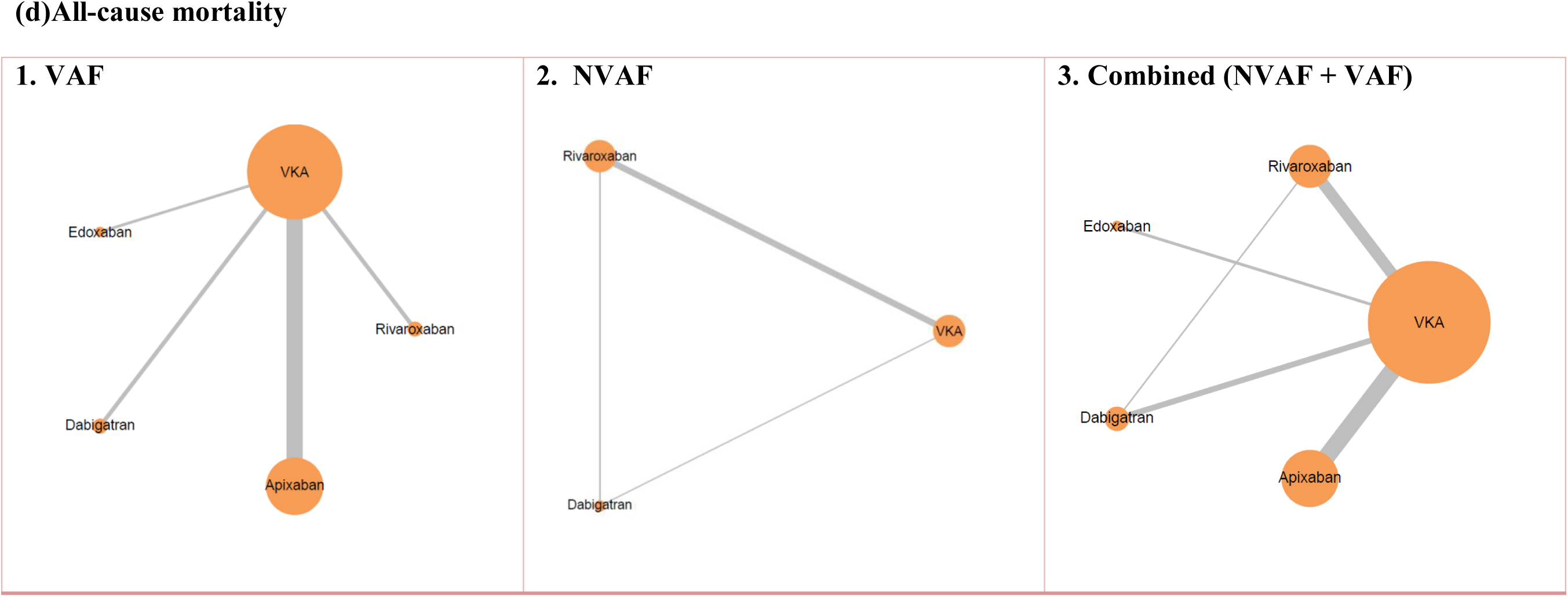
(a-d): Network plots illustrating the evidence structure for outcomes: (a) ischemic stroke or systemic embolism (IS/SE), (b) hemorrhagic stroke (HS), (c) major bleeding, and (d) all-cause mortality. Each plot displays treatment comparisons across three population categories: (a) VAF, (b) NVAF, and (c) combined (NVAF + VAF).

### Main Findings

#### Ischemic Stroke (IS) or Systemic Embolism (SE)

In the overall analysis, DOACs showed a favorable effect compared to VKAs in reducing the risk of IS or SE. Apixaban (OR: 0.81, 95% CrI: 0.73 to 0.90), dabigatran (OR: 0.77, 95% CrI: 0.68 to 0.87), and rivaroxaban (OR: 0.87, 95% CrI: 0.79 to 0.96) demonstrated significant risk reduction, whereas edoxaban (OR: 1.05, 95% CrI: 0.89 to 1.22) did not show a statistically significant effect. In subgroup analysis, no DOAC showed a significant effect in NVAF patients, including dabigatran (OR: 0.77, 95% CrI: 0.50 to 1.19) and rivaroxaban (OR: 0.86, 95% CrI: 0.70 to 1.02). In contrast, VAF subgroup analysis revealed significant protection with apixaban (OR: 0.81, 95% CrI: 0.73 to 0.91) and dabigatran (OR: 0.77, 95% CrI: 0.66 to 0.90), while edoxaban and rivaroxaban showed no significant benefit.

#### Hemorrhagic Stroke (HS)

In the combined population, all four DOACs were associated with a significantly reduced risk of HS compared to VKAs: dabigatran (OR: 0.30, 95% CrI: 0.23 to 0.40), apixaban (OR: 0.53, 95% CrI: 0.42 to 0.66), edoxaban (OR: 0.47, 95% CrI: 0.35 to 0.63), and rivaroxaban (OR: 0.60, 95% CrI: 0.46 to 0.79). In NVAF, only dabigatran showed a significant reduction (OR: 0.28, 95% CrI: 0.08 to 0.92), while rivaroxaban was not significant. Among VAF patients, all DOACs demonstrated a significant reduction in HS risk.

#### Major Bleeding

Apixaban was the only DOAC associated with a significant reduction in major bleeding in the overall analysis (OR: 0.69, 95% CrI: 0.55 to 0.88). Dabigatran, edoxaban, and rivaroxaban did not show statistically significant differences. In the NVAF subgroup, neither dabigatran (OR: 0.79, 95% CrI: 0.21 to 2.66) nor rivaroxaban (OR: 1.21, 95% CrI: 0.70 to 1.99) showed significant effects. In contrast, among VAF patients, apixaban remained significantly protective (OR: 0.68, 95% CrI: 0.60 to 0.77), while the other DOACs showed non-significant trends toward reduced bleeding.

#### All-Cause Mortality

In the pooled analysis, apixaban (OR: 0.88, 95% CrI: 0.81 to 0.96) and dabigatran (OR: 0.89, 95% CrI: 0.81 to 0.99) were significantly associated with reduced all-cause mortality compared to VKAs. Edoxaban and rivaroxaban did not demonstrate significant associations. Among NVAF patients, apixaban showed a significant mortality benefit (OR: 0.88, 95% CrI: 0.82 to 0.96), while dabigatran, edoxaban, and rivaroxaban did not. In VAF, neither dabigatran (OR: 0.88, 95% CrI: 0.53 to 1.33) nor rivaroxaban (OR: 0.91, 95% CrI: 0.71 to 1.21) significantly reduced mortality.

#### Litmus Rank-O-Gram and SUCRA-Based Ranking

The Litmus Rank-O-Gram SUCRA plots [Figures 4 (a–d)] visually summarize the probability-based rankings of treatment effectiveness across outcomes and subgroups. For IS/SE, dabigatran consistently ranked highest across all populations (combined, NVAF, VAF) with a SUCRA probability of 97%, indicating the greatest likelihood of being the most effective treatment. Apixaban and rivaroxaban followed, while edoxaban ranked lowest (5%), suggesting the least benefit in IS or SE prevention.

**Figure 3.**
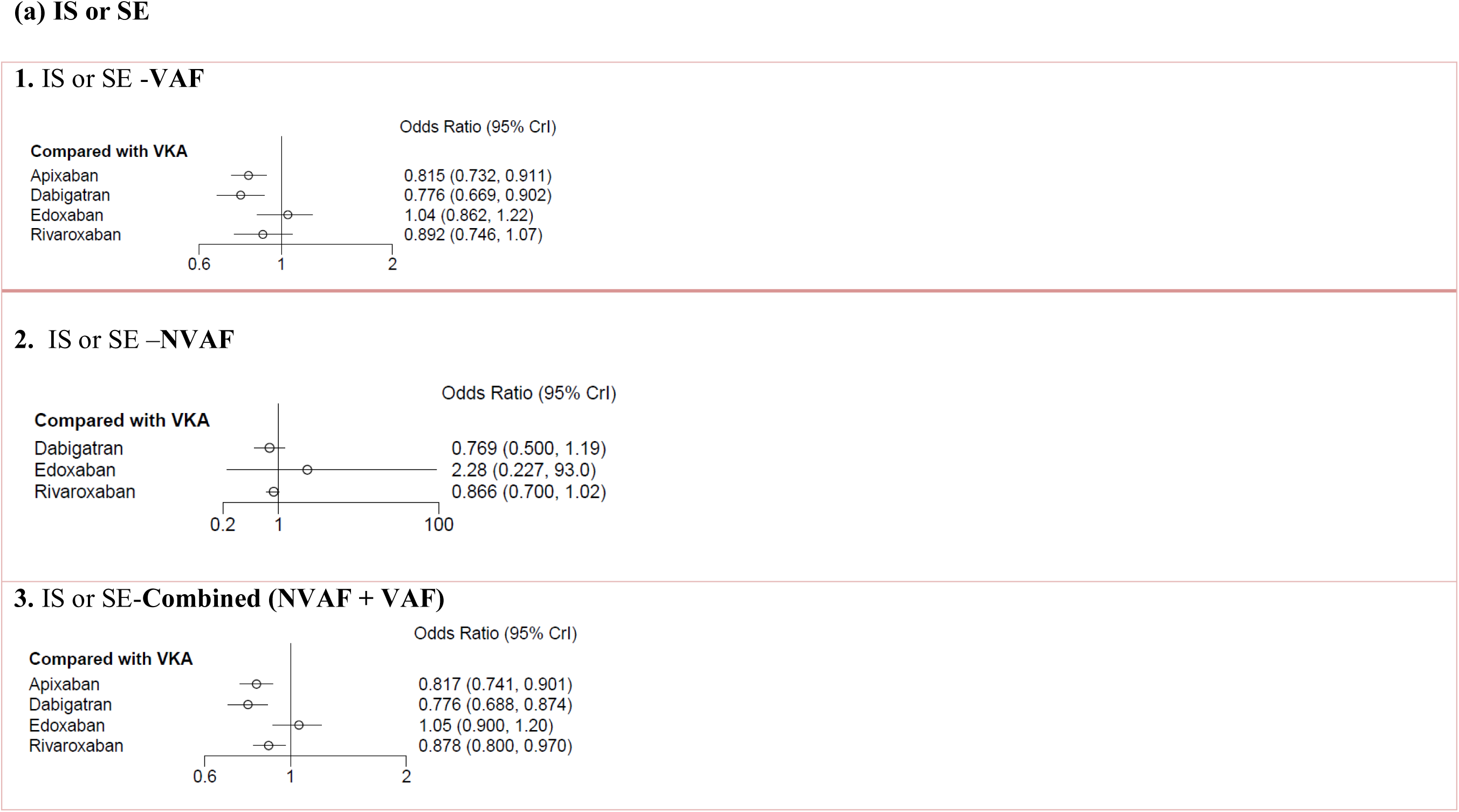
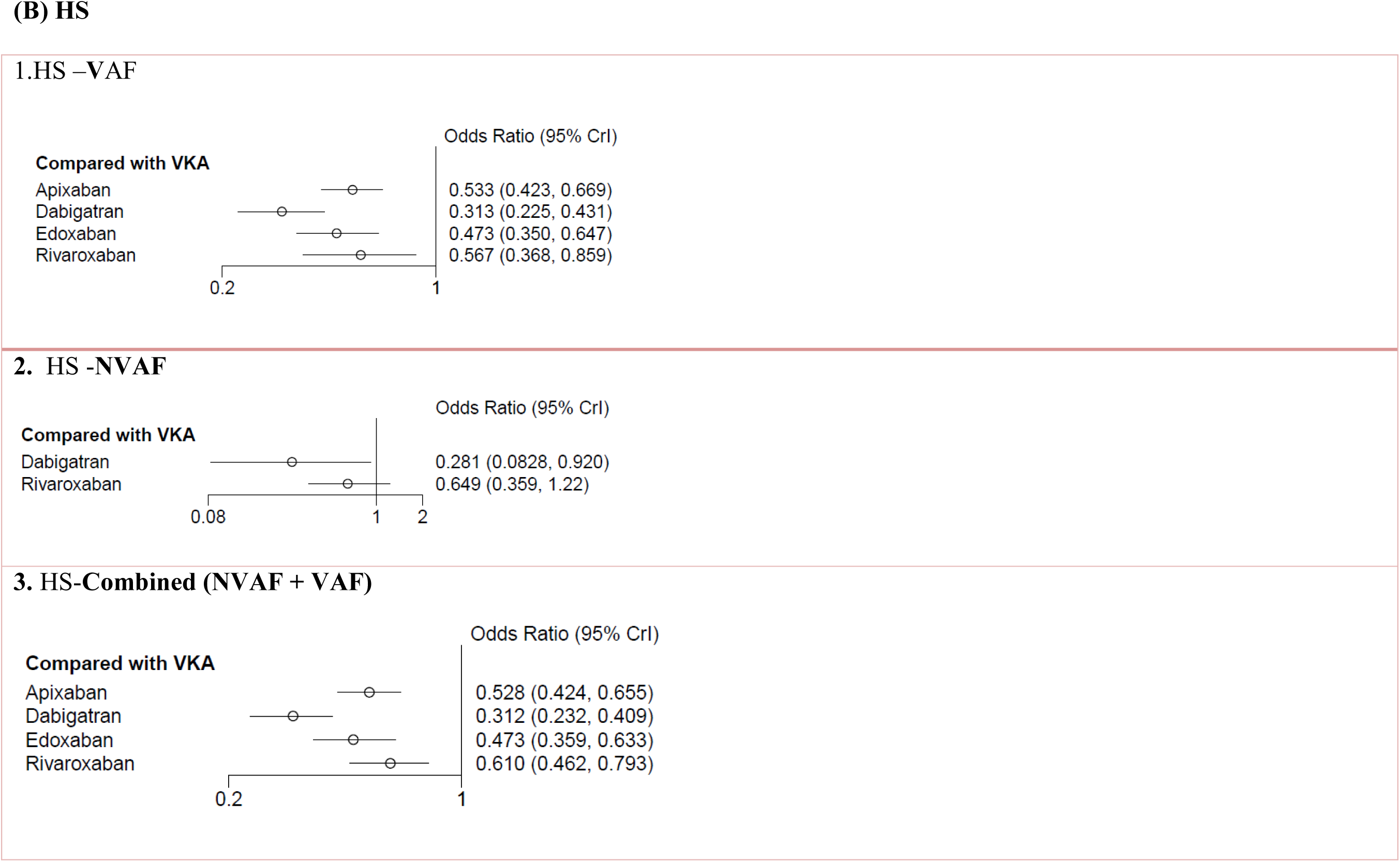
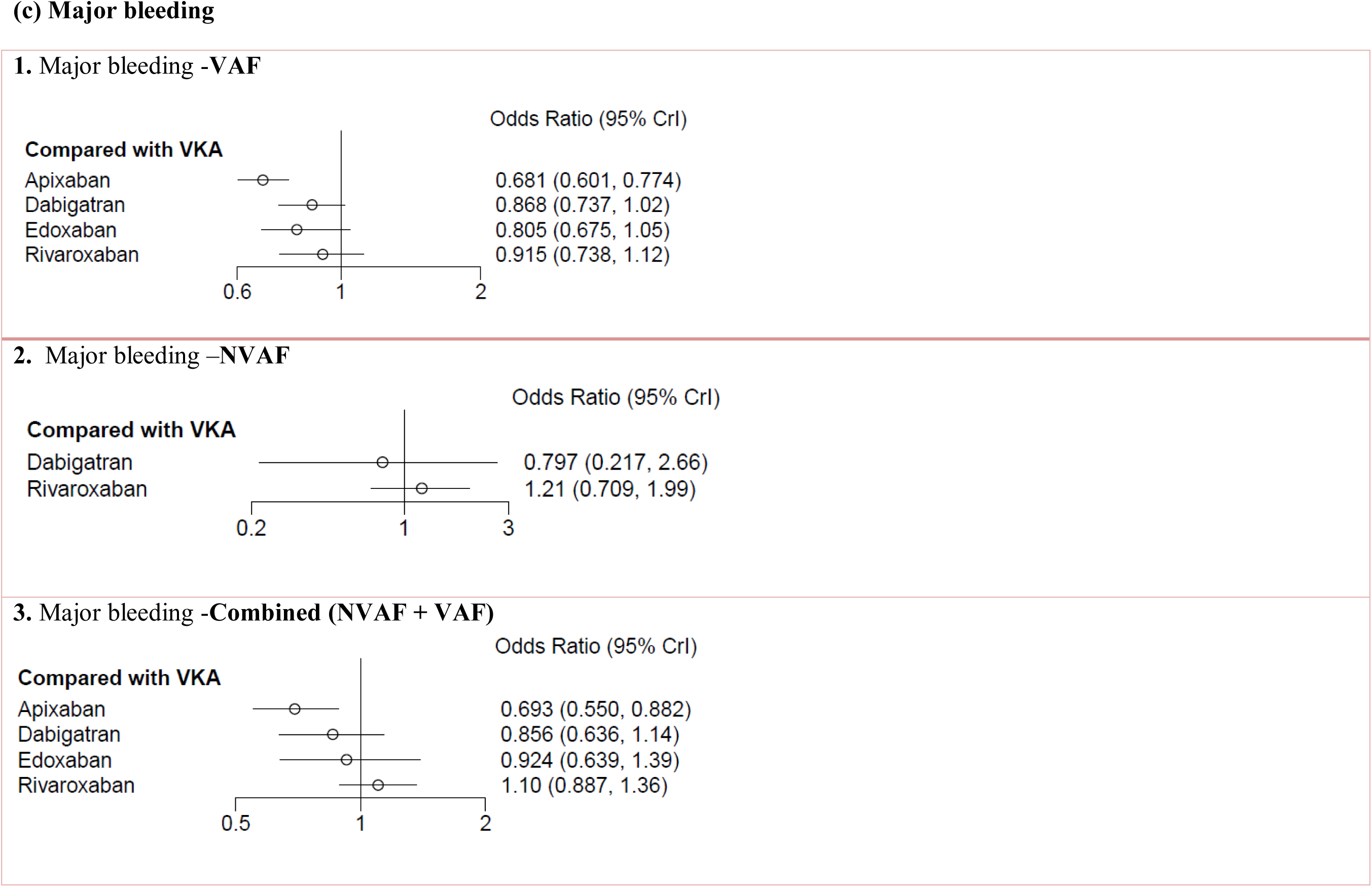
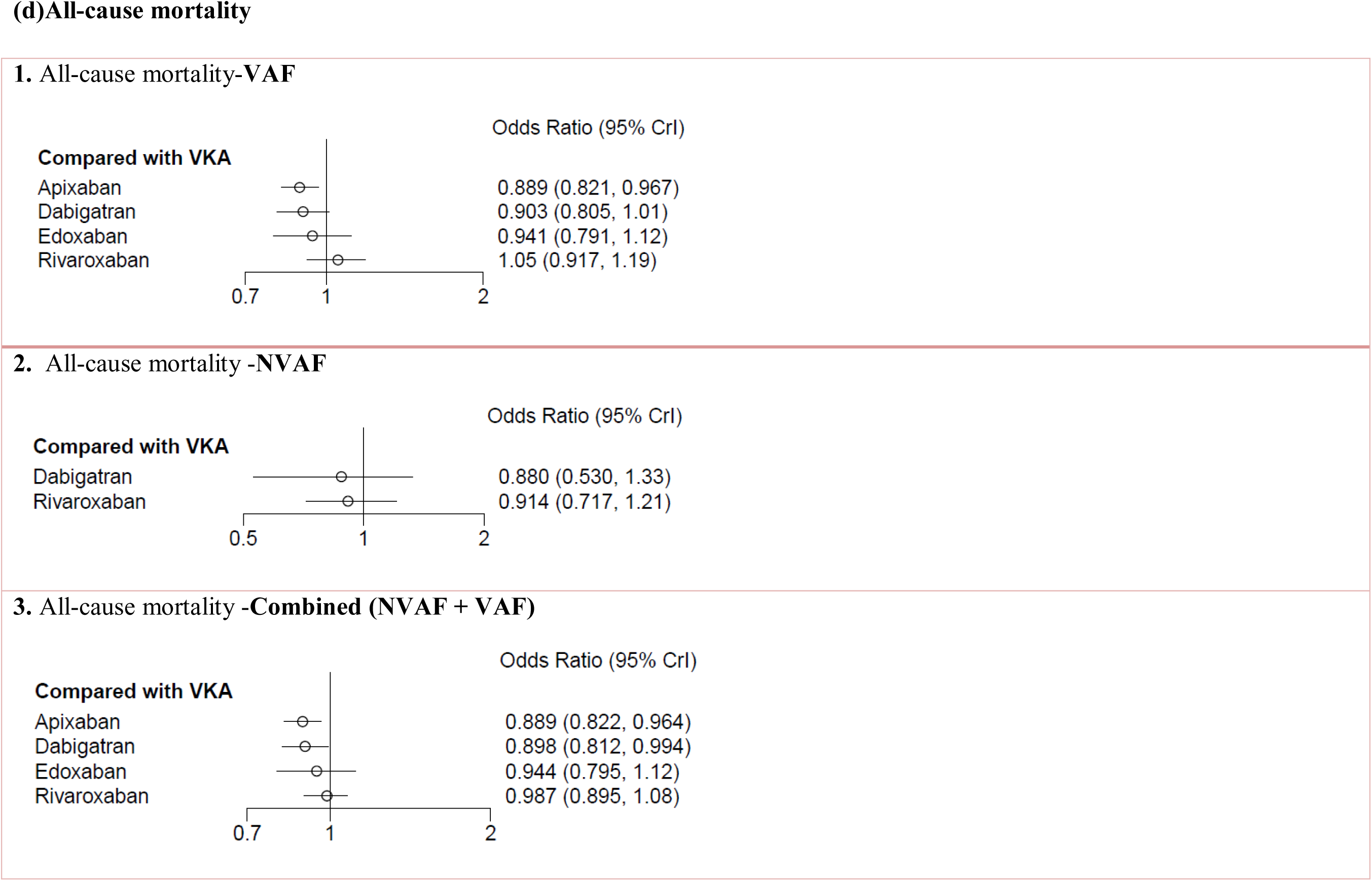
(a-d): Forest plots of relative treatment effects based on the Bayesian random-effects consistency model for: (a) ischemic stroke or systemic embolism (IS/SE), (b) hemorrhagic stroke (HS), (c) major bleeding, and (d) all-cause mortality. Each panel presents results stratified by: (1) VAF (2) NVAF, and (3) combined population (NVAF + VAF)

**Figure 4.**
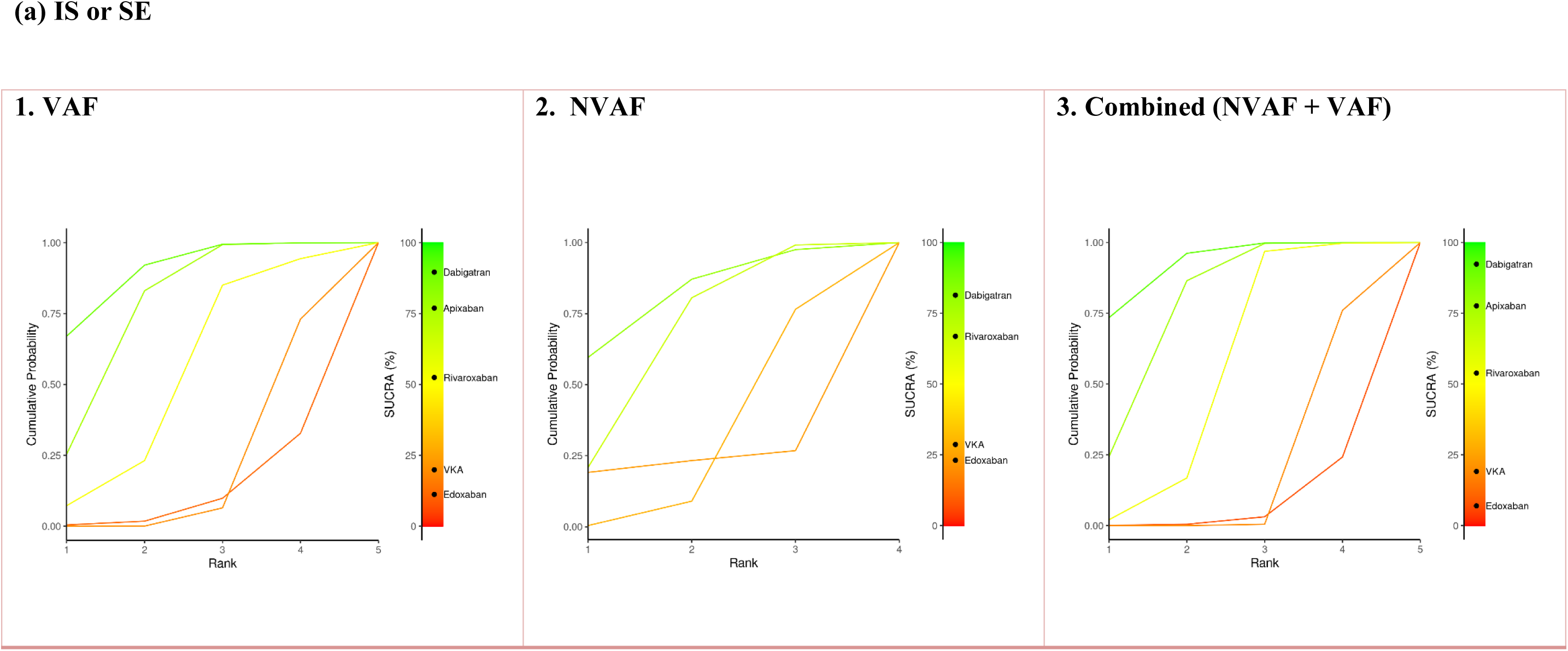
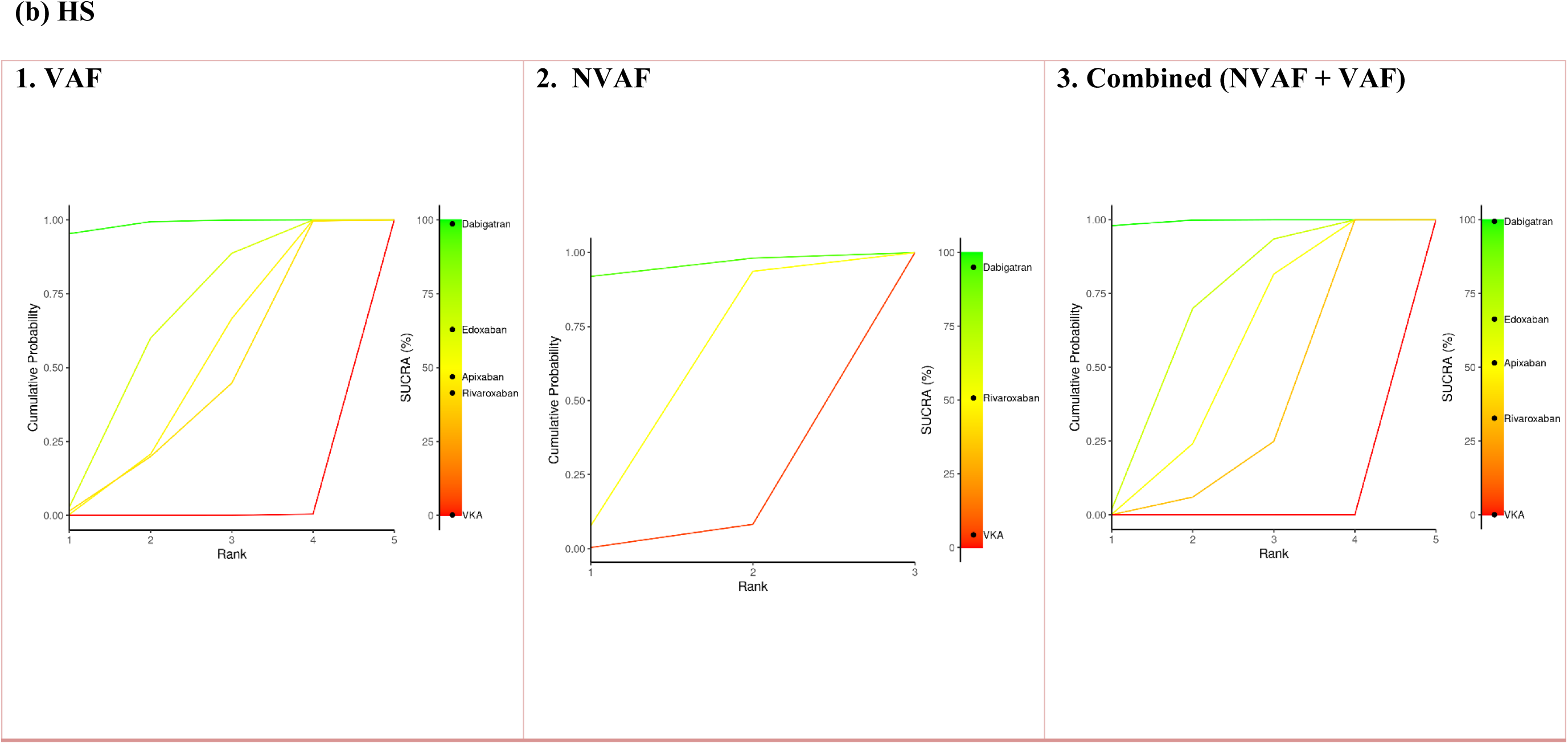
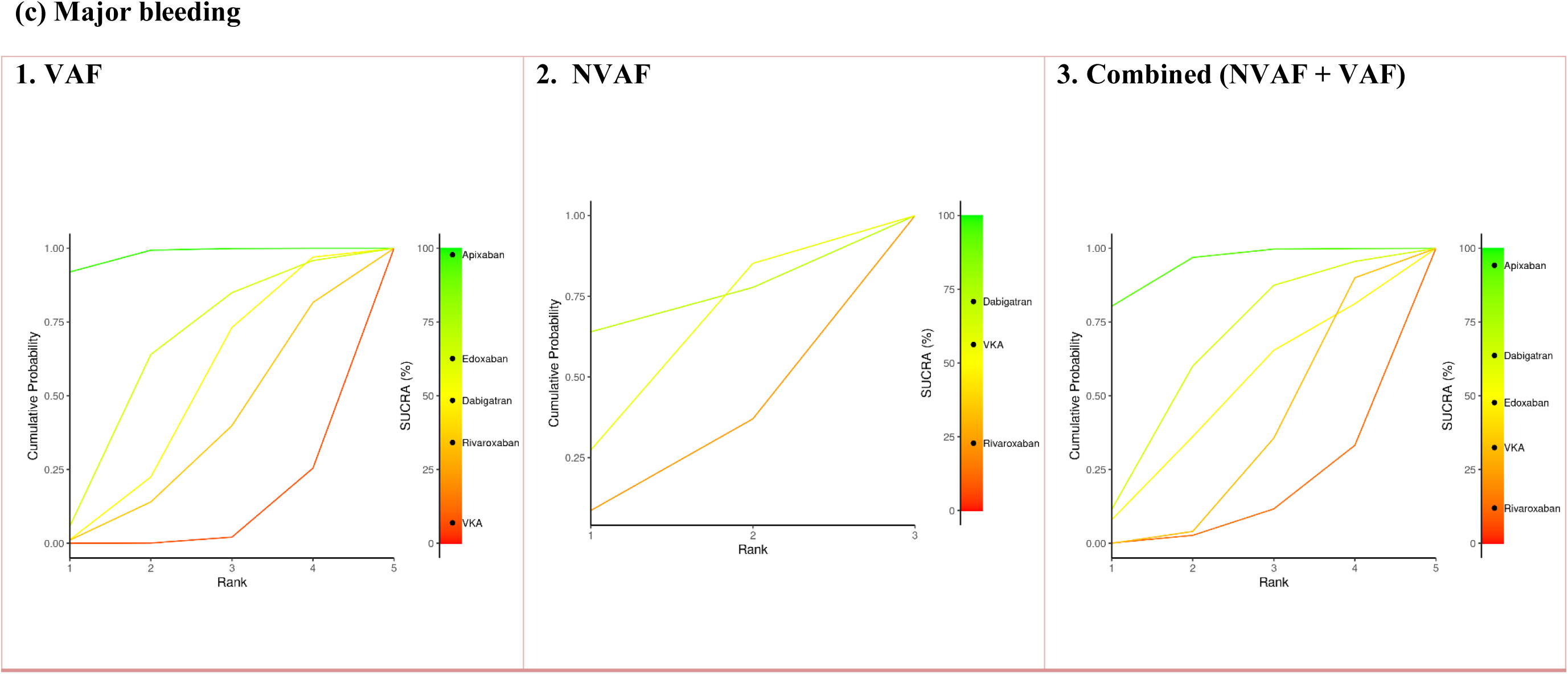
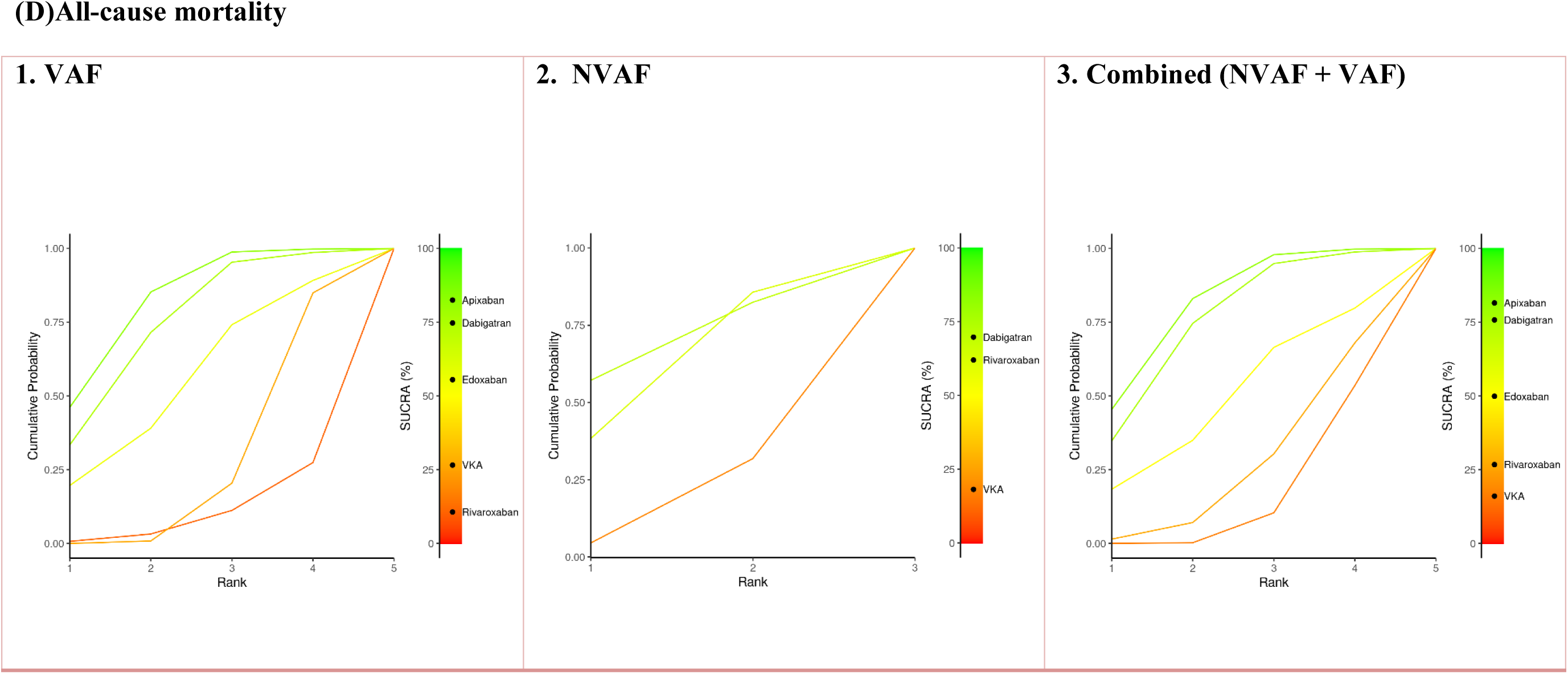
(a–d): Litmus-o-SUCRA plots displaying treatment rankings for: (A) ischemic stroke or systemic embolism (IS/SE), (B) hemorrhagic stroke (HS), (C) major bleeding, and (D) all-cause mortality. Rankings are shown across three categories: (1) VAF, (2) NVAF, and (3) combined population (NVAF + VAF).

**Table 1:** Baseline characteristics of included studies evaluating the efficacy of DOACs versus VKAs in stroke prevention among patients with AF.

| S.No. | Author & Year | Country | Study period | Inclusion criteria | Intervention (I) | Comparator (C) | Sample Size (I)/(C) | Mean Age (I) | Mean Age (C) | M/F (I) | M/F (C) | Follow up | Outcome measures | Quality Score |
| --- | --- | --- | --- | --- | --- | --- | --- | --- | --- | --- | --- | --- | --- | --- |
| 1. | Ezekowitz et al. 2007<sup>18</sup> | Denmark | NR | NVAF patients with CAD plus $\geq 1$ risk factor for stroke | Dabigatran 150 mg BD | Warfarin 5 mg OD | 166/70 | 70 ± 8.1 | 69 ± 8.3 | 135/31 | 59/11 | 12 weeks | MB | 7 |
| 2. | Connolly et al. 2009<sup>7</sup> | Canada | Dec. 2005 to Dec. 2007 | AF patients with increased risk for stroke | Dabigatran 110 mg BD<br>Dabigatran 150 mg BD | Warfarin 5 mg OD | 6015<br>6076/6022 | 71.4±8.6<br>71.5±8.8 | 71.6±8.6 | 3865/2150<br>3840/2236 | 3809/2213 | 2 years | IS or SE, MB, MI | 7 |
| 3. | Diener et al. 2010<sup>19</sup> | Germany | Dec. 2005 to Dec. 2007 | AF patients with 1 additional risk factor for stroke | Dabigatran 110 mg BD<br>Dabigatran 150 mg BD | Warfarin 5 mg OD | 4819<br>4843/4827 | 71.7 ± 8.4<br>71.7 ± 8.5 | 71.9±8.3 | 3099/1720<br>3073/1770 | 3063/1764 | 2 years | IS or SE, MB, MI death, MI, TIA | 5 |
| 4. | Weitz et al. 2010<sup>20</sup> | Canada | NR | NVAF patients with a CHADS2 score of at least 2 | Edoxaban 30 mg OD<br>Edoxaban 30 mg BD<br>Edoxaban 60 mg OD<br>Edoxaban 60 mg BD | Warfarin 5 mg OD | 235<br>244<br>234<br>180/250 | 65.2 ± 8.3<br>64.8 ± 8.8<br>64.9 ± 8.8<br>64.7 ± 9.0 | 66.0 ± 8.5 | 140/95<br>150/94<br>155/79<br>114/66 | 151/99 | NR | MB | 5 |
| 5. | Chung et al. 2011<sup>21</sup> | Korea | Oct. 2007 to Oct. 2008 | NVAF patients with a CHADS2 index score $>1$ | Edoxaban 30 mg OD<br>Edoxaban 60 mg OD | Warfarin 5 mg OD | 79<br>80/75 | 64.9 ± 9.1<br>65.9 ± 7.7 | 64.5 ± 9.5 | 51/28<br>55/25 | 47/28 | 12 weeks | MB | 6 |
| 6. | Eikelboom et al. 2011<sup>22</sup> | Canada | NR | AF patients who had at least 1 additional risk factor for stroke | Dabigatran 110 mg BD<br>Dabigatran 150 mg BD | Warfarin 5 mg OD | 6015<br>6076/6022 | 71.4±8.6<br>71.5±8.8 | 71.6±8.6 | 3865/2150<br>3840/2236 | 3809/2213 | 2 years | IS or SE, MB | 4 |
| 7. | Fox et al. 2011<sup>23</sup> | France | NR | NVAF patients with moderate-to-high risk of stroke | Rivaroxaban 15 mg OD<br>Rivaroxaban 20 mg OD | Warfarin 5 mg OD | 1474<br>5637/5640 | 78.75±1.75<br>70.25±2.16 | 70.25±2.16 | 663/811<br>3629/2008 | 3641/1999 | NR | IS or SE, MB, MI death, MI | 6 |
| 8. | Granger et al. 2011<sup>6</sup> | UK | Dec. 2006 to Apr. 2010 | AF patients with at least 1 additional risk factor for stroke | Apixaban 5 mg BD | Warfarin 5 mg OD | 9120/9081 | 69.75±2.16 | 69.75±2.16 | 5886/3234 | 5899/3182 | 2 years | IS or SE, MB, MI death, MI, HS | 7 |
| 9. | Patel et al. 2011<sup>5</sup> | UK | Dec. 2006 to May 2010 | NVAF patients with moderate-to-high risk for stroke | Rivaroxaban 20 mg OD | Warfarin 5 mg OD | 7131/7133 | 72.25±2.16 | 72.25±2.16 | 4300/2831 | 4301/2832 | 707 days | IS or HS and SE, death, MI | 8 |
| 10. | Easton et al. 2012<sup>24</sup> | USA | Dec 2006 to April 2010 | AF patients who were randomly assigned | Apixaban 5 mg BD | Warfarin 5 mg OD | 7426/7339 | NR | NR | NR | NR | 1.8 years | Stroke/SE, MB | 7 |
| 11. | Hankey et al. 2012<sup>25</sup> | UK | Dec. 2006 to Jun. 2009 | AF patients with 1 additional risk factor for stroke | Rivaroxaban 20 mg OD | Warfarin 5 mg OD | 3377/3419 | 74.25±1.83 | 74±2 | 2029/1348 | 2037/1382 | 676 days | IS or SE, MB, MI death, MI, TIA, ICH | 7 |
| 12. | Hori et al. 2012<sup>26</sup> | Japan | June 2007 to Jan. 2010 | NVAF patients with at least 1 additional risk factor for stroke | Rivaroxaban 15 mg OD | Warfarin 5 mg OD | 639/639 | 71±13.7 | 71.2±11.7 | 530/109 | 500/139 | 30 days | IS or SE, MB, MI death | 8 |
| 13. | Huisman et al. 2012<sup>27</sup> | Netherlands | NR | AF patients who had differing risks of MI and were treated with different comparators | Dabigatran 110 mg BD<br>Dabigatran 150 mg BD | Warfarin 5 mg OD | 6015<br>6076/6022 | NR | NR | NR | NR | 12 month | Stroke/SE, HS, MI, death, MB | 4 |
| 14. | Yamashita et al. 2012<sup>28</sup> | Japan | Apr 2007 to July 2008 | NVAF patients CHADS2 score ≥1 | Edoxaban 30 mg OD<br>Edoxaban 45 mg BD<br>Edoxaban 60 mg OD | Warfarin 5 mg OD | 131<br>134<br>131/129 | 69.4<br>69.5<br>68.4 | 68.8 | 110/21<br>109/25<br>107/24 | 107/22 | 12 weeks | ICH, MB | 6 |
| 15. | Al-Khatib et al. 2013<sup>29</sup> | USA | NR | AF patients with at least one risk factors for stroke | Apixaban 5 mg BD | Warfarin 5 mg OD | 7744/7668 | NR | NR | NR | NR | NR | IS/HS/SE, death | 5 |
| 16. | Bahit et al. 2013<sup>30</sup> | USA | 2006 to 2010 | AF patients ≥80 years | Apixaban 5 mg BD | Warfarin 5 mg OD | 1316/1512 | NR | NR | NR | Nr | 1.8 years | IS or SE, MB, death, MI | 4 |
| 17. | Diepen et al. 2013<sup>31</sup> | Canada | NR | NVAF patients with moderate to high risk of stroke | Rivaroxaban 20 mg OD | Warfarin 5 mg OD | 2551/2587 | 73.25±2.16 | 73.25±1.83 | 1522/1029 | 1544/1043 | NR | IS or HS, SE, death, MI, MB | 5 |
| 18. | Flaker et al. 2013<sup>32</sup> | Columbia | NR | AF patients with at least 1 of the risk factors for stroke | Apixaban 5 or 2.5 mg BD | Warfarin 5 mg OD | 265/275 | 67.1 (9.2) | 67.3 (9.4) | 193/72 | 201/74 | 30 days | IS/SE, MB, MI death, MI | 5 |
| 19. | Garcia et al. 2013<sup>33</sup> | USA | NR | AF patients with 1 risk factor for stroke | Apixaban 5 mg BD | Warfarin 5 mg OD | 3912/3888 | 70 (62, 76) | 70 (62, 76) | 2398/1514 | 2363/1525 | NR | Stroke/SE, MB | 6 |
| 20. | Giugliano et al. 2013<sup>8</sup> | USA | 19 Nov. 2008 to 22 Nov. 2010 | AF patients with CHADS<sub>2</sub> score of ≥2 | Edoxaban 60 mg OD<br>Edoxaban 30 mg OD | Warfarin 5 mg OD | 7035<br>7034/7036 | 74±0.6<br>74±0.6 | 74±0.6 | 4366/2669<br>4304/2730 | 4395/2641 | 2.8 years | Stroke, SE, MB, death | 8 |
| 21. | Mahaffey et al. 2013<sup>34</sup> | UK | 18 Dec. 2006 to 17 June 2009 | AF patients with VKA therapy | Rivaroxaban 20 mg OD | Warfarin 5 mg OD | 3175/3192 | 70.3±9.8 | 70.4 ±9.5 | 1799/1376 | 1813/1379 | NR | Stroke, SE, MB | 5 |
| 22. | Tanahashi et al. 2013<sup>35</sup> | Japan | NR | NVAF patients with 1 additional risk factor for stroke | Rivaroxaban 15 mg OD | Warfarin 5 mg OD | 231/234 | 72.2±8.28 | 72.6±8.46 | 185/46 | 173/61 | 30 month | Stroke, SE, MB | 6 |
| 23. | Alexander et al. 2014<sup>36</sup> | USA | NR | AF patients with at least 1 additional risk factor for stroke | Apixaban 5 mg BD | Warfarin 5 mg OD | 6852/6847 | NR | NR | NR | NR | 1.8 years | IS/SE, MB, MI Death | 5 |
| 24. | Breithardt et al. 2014<sup>37</sup> | Germany | NR | NVAF patients who, artificial heart valves repair | Rivaroxaban 20 mg OD | Warfarin 5 mg OD | 968/1035 | NR | NR | NR | NR | 3 years | IS/SE, MB, MI Death | 5 |
| 25. | Cappato et al. 2014<sup>38</sup> | Italy | NR | NVAF patients with naïve or experienced OAC with VKA | Rivaroxaban 20 mg OD | VKA 5 mg OD | 585/287 | 65.3±10.4 | 65.3±10.6 | 426/159 | 203/84 | 30 days | IS/SE, MB, MI death, MI, TIA | 4 |
| 26. | Halperin et al. 2014<sup>39</sup> | USA | NR | NVAF patients with 1 additional risk factor for stroke | Rivaroxaban 15 mg OD | Warfarin 5 mg OD | 4011/4024 | 65.75±1.83 | 65.75±1.83 | 2627/1384 | 2626/1398 | 707 days | IS or SE, MB, MI death | 7 |
| 27. | Mao et al. 2014<sup>40</sup> | China | July 2009 to Dec. 2010 | AF patients with 1 additional risk factor for stroke | Rivaroxaban 20 mg OD | Warfarin 5 mg OD | 177/176 | 74.25±1.83 | 74.25±1.83 | 198/69 | 110/66 | NR | IS or SE, MB, MI death | 6 |
| 28. | Avezum et al. 2015<sup>41</sup> | Brazil | NR | AF patients with 1 additional risk factor for stroke | Apixaban 5 mg BD | Warfarin 5 mg OD | 2438/2370 | NR | NR | NR | NR | NR | IS or SE, MB, MI death | 3 |
| 29. | Duraes et al. 2016<sup>42</sup> | Brazil | Aug. 2013 to Nov. 2014 | AF patients with aortic bioprostheses valve replacement | Dabigatran 110 mg BD | Warfarin 5 mg OD | 15/12 | 48.8±10.4 | 45.7±6 | 5/10 | 5/7 | 90 days | IS or SE, MB, MI death | 4 |
| 30. | Gibson et al. 2016<sup>43</sup> | USA | May 2013 to July 2015 | AF patients who had receiving OAC for 3 months immediately | Rivaroxaban 15 mg OD | Warfarin 5 mg OD | 709/706 | 70.4±9.1 | 69.9±8.7 | 528/181 | 518/188 | 12 months | MB | 6 |
| 31. | Goette et al. 2016<sup>44</sup> | Germany | Mar. 2014 to Oct. 2015 | NVAF patients who planned for electrical cardioversion n | Edoxaban 60 mg OD | Warfarin 5 mg OD | 506/510 | NR | NR | 336/170 | 333/177 | 28 days | IS or SE, MB, MI death | 8 |
| 32. | Rost et al. 2016<sup>45</sup> | USA | NR | AF patients with planned anticoagulation therapy | Edoxaban 60 mg OD | Warfarin 5 mg OD | 5059/5045 | NR | NR | NR | NR | 2.8 years | Stroke/SE | 6 |
| 33. | Lauw et al. 2017<sup>46</sup> | Canada | NR | AF patients with at least 1 additional risk factor for stroke | Dabigatran 110 mg BD<br>Dabigatran 150 mg BD | Warfarin 5 mg OD | 6015<br>6076/6022 | NR | NR | NR | NR | 3 years | Stroke/SE, MB | 3 |
| 34. | Ezekowitz et al. 2018<sup>47</sup> | USA | July 2014 to Feb. 2017 | AF patients with < 48 h of prior anticoagulation | Apixaban 5 mg BD | Warfarin 5 mg OD | 753/747 | 64.7 ± 12.2 | 64.5 ± 12.8 | 505/248 | 497/250 |  | Stroke, SE, all-cause death | 6 |
| 35. | Kimura et al. 2018<sup>48</sup> | Japan | July 2014 to Jan. 2016 | NVAF patients with left atrial diameter of ≥55 mm | Rivaroxaban 20 mg OD | Warfarin 5 mg OD | 64/63 | NR | NR | 53/11 | 53/10 |  | Thromboembolism and MB | 7 |
| 36. | Kirchhof et al. 2018<sup>49</sup> | UK |  | AF patients with at least 1 stroke risk factor | Apixaban 5 mg BD | Warfarin 5 mg OD | 318/315 | NR | NR | 218/100 | 206/109 | 3 months | Stroke, bleeding, death | 7 |
| 37. | Guimaraes et al. 2019<sup>50</sup> | USA | NR | AF patients with ≥1 risk factors for stroke | Apixaban 5 mg BD | Warfarin 5 mg OD | 87/69 | NR | NR | 53/34 | 42/27 | NR | IS or SE, MB, MI death | 4 |
| 38. | Vranckx et al. 2019<sup>51</sup> | Belgium | Feb 2017 to May 2018 | AF patients had a successful PCI for stable coronary artery disease | Edoxaban 30 mg OD | VKA 5 mg OD | 751/755 | 69 (63–77) | 70 (64–77) | 557/194 | 563/192 | NR | IS or SE | 6 |
| 39. | Guimaraes et al. 2020<sup>52</sup> | Brazil | 14 April 2007 to 22 July 2019 | AF patients with persistent bioprosthetic mitral valve | Rivaroxaban 20 mg OD | Warfarin 5 mg OD | 500/505 | 59.4±2.4 | 59.2±11.8 | 189/311 | 209/296 | 12 months | MB | 7 |
| 40. | Mieghem et al. 2021<sup>53</sup> | USA | Apr. 2017 to Jan. 2020 | AF patients after transcatheter aortic-valve replacement | Edoxaban 60 mg OD | VKA 5 mg OD | 713/713 | 82.1±5.4 | 82.1±5.5 | 366/347 | 382/331 | 36 months | IS, SE, MI, MB, death | 5 |
| 41. | Ahasan et al. 2022<sup>54</sup> | Dhaka | Nov. 2019 to Oct. 2020 | NVAF patients with consent | Dabigatran 110 mg BD | Rivaroxaban 20 mg OD | 37/37 | 59.92±10.13 | 61.42±11.21 | 24/13 | 21/16 | 6 months | IS, bleeding | 4 |
| 42. | Connolly et al. 2022<sup>55</sup> | Canada |  | AF patient with CHA<sub>2</sub>DS<sub>2</sub>V ASc score of at least 2 | Rivaroxaban 20 mg OD | VKA 5 mg OD | 2275/2256 | 50.7±14.8 | 50.3±14.4 | 627/1648 | 630/2256 | 3.1 years | Stroke, SE, MI or death | 7 |
| 43. | Pokorney et al. 2022<sup>56</sup> | UK | Jan. 2017 to Jan. 2019 | AF patients with at least 1 of stroke prevention factors | Apixaban 5 mg BD | Warfarin 5 mg OD | 82/72 | NR | NR | 48/34 | 50/22 | 340 days | Stroke, death, bleeding | 4 |

RCT= Randomized control trial, RCS= Retrospective cohort study, PCS= Prospective cohort study, OD= Once a day, BD= Twice a day, NR= Not reported, IS= Ischemic stroke, SE= Systemic embolism, MI= Myocardial infarction, TIA= Transient ischemic attack, HS= Hemorrhagic stroke, Electrocardiography, AF= Atrial fibrillation, NVAF= Non-valvular atrial fibrillation, ICH= Intracranial bleeding, GIB= Gastrointestinal bleeding, MB= Major bleeding, ACM= all-cause mortality, CRNM= Clinically relevant non-major, NMCR= Non-major clinically relevant bleeding, TE= Thromboembolism, VTE= Venous thromboembolism, DVT= deep-vein thrombosis, PE= Pulmonary embolism, OACs= Oral anticoagulants, NOACs= Non-vitamin K antagonist OACs, ICD-9= International Classification of Diseases, Ninth Revision, CHA<sub>2</sub>DS<sub>2</sub>-VAsC = congestive heart failure, hypertension, age 75 years or older, diabetes mellitus, previous stroke/transient ischemic attack, vascular disease, age 65 to 74 years, female, CAD= Coronary artery disease

For HS, dabigatran again achieved the highest SUCRA score (98%) across all subgroups. Notably, rivaroxaban performed better in the NVAF subgroup but was less effective in the combined and VAF populations. In the context of major bleeding, apixaban ranked highest in the combined and VAF subgroups (97%), whereas dabigatran ranked highest (73%) in the NVAF group. Regarding all-cause mortality, apixaban (78%) and dabigatran (74%) emerged as the most effective treatments in the combined and VAF populations. In the NVAF subgroup, dabigatran led with a SUCRA value of 73%, suggesting the greatest probability of reducing mortality.

#### Quality Assessment

All 43 included RCTs were assessed for methodological quality using standardized risk-of-bias tools and were determined to have a **low risk of bias** (Table S2). The rigorous evaluation ensured the internal validity and reliability of the synthesized evidence. Moreover, the overall consistency and homogeneity observed across studies further strengthen the credibility and robustness of the network meta-analysis findings.

## Discussion

This network meta-analysis synthesizes evidence from 43 RCTs comparing the effectiveness and safety of four DOACs—dabigatran, apixaban, rivaroxaban, and edoxaban—versus VKAs or placebo across clinically relevant outcomes in patients with AF. By incorporating both direct and indirect comparisons and evaluating treatment effects across combined, NVAF, and VAF populations, our study offers a comprehensive and updated assessment of the comparative performance of these anticoagulants in routine clinical practice.

Our findings reinforce the clinical value of DOACs, particularly dabigatran and apixaban, for the prevention of thromboembolic events. Dabigatran consistently ranked highest in preventing IS or SE across all subgroups, supported by robust SUCRA values (97%) and statistically significant effect estimates. This observation aligns with the RE-LY trial^7^ and earlier NMAs, such as that by López-López et al. (2017),^57^ which demonstrated superior stroke prevention with dabigatran relative to VKAs and other DOACs. Apixaban and rivaroxaban also showed favorable efficacy, although their rank probabilities and effect sizes were slightly lower. Notably, edoxaban consistently ranked lowest, suggesting comparatively reduced efficacy—a finding that contrasts with some earlier pooled analyses (e.g., Ruff et al., 2014^58^) that reported broadly similar efficacy across DOACs. The present analysis, enriched with updated trial data and subgroup stratification, highlights meaningful inter-agent variability that may influence therapeutic decision-making.

For HS, all four DOACs demonstrated statistically significant risk reduction compared to VKAs, with dabigatran again achieving the highest SUCRA ranking (98%). These findings are concordant with prior studies indicating that DOACs, as a class, are associated with lower risk of intracranial bleeding than VKAs. However, our stratified analysis revealed important nuances— rivaroxaban was notably more protective in NVAF but less so in VAF or the overall population. This subgroup-specific variation may relate to differing patient characteristics, comorbidities, or procedural factors such as valvular pathology or concomitant antiplatelet use, and underscores the need for individualized risk assessment.

With respect to major bleeding, apixaban emerged as the most favorable agent in the combined and VAF populations, consistent with the ARISTOTLE trial^7^ and meta-analyses by Caldeira et al. (2015)^59^ and Cohen et al. (2016),^60^ which emphasized apixaban’s bleeding advantage among the DOACs. In contrast, dabigatran ranked higher for bleeding safety in NVAF patients, albeit with less consistent statistical significance. Rivaroxaban and edoxaban exhibited neutral or slightly increased bleeding risk in various subgroups. These findings support the view that not all DOACs are interchangeable in safety profiles and that agent selection must consider individual bleeding risk, renal function, and patient adherence.

In terms of all-cause mortality, apixaban and dabigatran again showed the greatest benefit. Apixaban ranked highest in the overall and VAF groups, while dabigatran was most favorable in NVAF. Although mortality outcomes are often influenced by multiple factors beyond anticoagulant therapy, our findings suggest that these agents offer a favorable balance of stroke prevention and safety that may translate into improved survival. This aligns with prior NMAs (e.g.,; Ntaios et al., 2020^61^, Chan et al. 2024^62^), though those analyses often lacked granularity regarding subgroups or excluded VAF populations entirely.

Another notable strength of this NMA is its application of both SUCRA and Litmus Rank-O-Gram approaches, which provide probabilistic insight into treatment hierarchies across multiple outcomes. The consistency of ranking for dabigatran and apixaban across both efficacy and safety endpoints highlights their favorable net clinical benefit and supports their use as preferred agents in most AF scenarios. Conversely, the lower rankings of edoxaban raise questions about its comparative role in long-term stroke prevention, particularly in high-risk or VAF populations.

### Limitations

Despite its strengths, this study has several limitations. First, although we included only RCTs to minimize confounding, variability in trial design, patient characteristics, and definitions of outcomes (e.g., major bleeding or HS) may have introduced heterogeneity. We attempted to address this by conducting subgroup analyses for NVAF and VAF, but some categories still lacked consistent or granular reporting. Second, the classification of AF as NVAF or VAF was not uniform across all studies. Some trials excluded patients with mechanical heart valves but included those with other valvular pathologies, potentially blurring subgroup distinctions. This heterogeneity in definitions may have influenced subgroup-specific findings and warrants caution in interpretation.

Third, although our NMA design allows integration of both direct and indirect evidence, it assumes consistency across comparisons and similar baseline risk across populations. While statistical consistency was acceptable, clinical heterogeneity cannot be entirely ruled out. Fourth, we did not include individual patient data, which limits our ability to assess treatment interactions by age, sex, renal function, or comorbidity burden—factors that are highly relevant in anticoagulant decision-making. Access to patient-level data would have allowed for more refined subgroup analyses and adjustment for potential effect modifiers. Fifth, our analysis did not distinguish between different dosing regimens or geographic variations in treatment protocols, both of which could influence outcome measures, particularly bleeding risk. Similarly, adherence and persistence, which are known to impact DOAC effectiveness in real-world settings, were not captured in RCT-level data. Finally, while we employed a comprehensive search strategy and rigorous quality assessment (with all included trials rated as low risk of bias), we cannot exclude the possibility of publication bias, particularly for more recently introduced agents or underreported adverse events.

## Conclusion

This updated and stratified NMA confirms that dabigatran and apixaban offer the most favorable efficacy-safety balance among currently available DOACs for stroke prevention in AF. These findings align with, and extend, previous meta-analytic evidence by incorporating a broader evidence base, applying population-specific comparisons, and using rank-based probabilistic approaches. The results support their use as first-line agents, particularly in patients with NVAF and VAF where individual risks for stroke, bleeding, and mortality must be carefully weighed. Further research through head-to-head RCTs and real-world data analyses is essential to validate these comparative advantages and guide personalized anticoagulant therapy.

## Data Availability

All the data and materials used in this systematic review and network meta-analysis are included in the manuscript. These contain detailed information on the datasets, including references to the original sources cited in the manuscript.

## Legend toSupplementary Tables

**Table-S1:** Summary of Search Results across Databases with Search Key Terms and results

**Table S2:** Quality assessment using a Modified Jadad Scoring of all the included RCTs

