## Supplementary material for "Direct Oral Anticoagulants for Stroke Prevention in Atrial Fibrillation: A Global Synthesis of Randomized Evidence through Network Meta-Analysis": NA

### Legend toSupplementary Tables

**Table-S1:** Summary of Search Results across Databases with Search Key Terms and results

**Table S2:** Quality assessment using a Modified Jadad Scoring of all the included RCTs

**Table-S1:** Summary of Search Results across Databases with Search Key Terms and results

| **Databases** | **Search Key Terms** | **Search Results** |
|---|---|---|
| PubMed | (("oral anticoagulants") OR ("anticoagulants") OR ("Direct oral anticoagulant") OR ("DOACs") OR ("different anticoagulants") OR ("new oral anticoagulants") OR ("NOACs") OR ("anticoagulant treatment") OR ("Dabigatran") OR ("Apixaban") OR ("Rivaroxaban") OR ("Edoxaban") OR ("Vitamin K antagonist") OR ("VKAs") OR ("warfarin")) AND (("stroke") OR ("stroke prevention") OR ("ischemic stroke") OR ("ischaemic stroke") OR ("hemorrhagic stroke") OR ("Thromboembolism") OR ("Systemic embolism")) AND (("atrial fibrillation") OR ("non-valvular atrial fibrillation")) AND (("heart disease") OR ("myocardial infarction")) AND (("major bleeding") OR ("minor bleeding") OR ("gastrointestinal bleeding") OR ("intracranial bleeding") OR ("all-cause mortality")) AND (("comparative effectiveness") OR ("safety effectiveness")) | 33 |
| EMbase | (oral anticoagulants OR anticoagulants OR "Direct oral anticoagulant" OR DOACs OR "different anticoagulants" OR "new oral anticoagulants" OR NOACs OR "anticoagulant treatment" OR Dabigatran OR Apixaban OR Rivaroxaban OR Edoxaban OR "Vitamin K antagonist" OR VKAs OR warfarin) AND (stroke OR "stroke prevention" OR "prevention of stroke" OR "prevention of cerebrovascular disorder" OR "ischemic stroke" OR "ischaemic stroke" OR "hemorrhagic stroke" OR Thromboembolism OR "Systemic embolism") AND (atrial fibrillation OR "non-valvular atrial fibrillation") AND ("heart disease" OR "myocardial infarction") AND ("major bleeding" OR "minor bleeding" OR "gastrointestinal bleeding" OR "intracranial bleeding" OR "all-cause mortality") AND ("comparative effectiveness" OR "safety effectiveness") | 798 |
| Cochrane Library | (("oral anticoagulants") OR ("anticoagulants") OR ("Direct oral anticoagulant") OR ("DOACs") OR ("different anticoagulants") OR ("new oral anticoagulants") OR ("NOACs") OR ("anticoagulant treatment") OR ("Dabigatran") OR ("Apixaban") OR ("Rivaroxaban") OR ("Edoxaban") OR ("Vitamin K antagonist") OR ("VKAs") OR ("warfarin")) AND (("stroke") OR ("stroke prevention") OR ("prevention of stroke") OR ("prevention of cerebrovascular disorder") OR ("ischemic stroke") OR ("ischaemic stroke") OR ("hemorrhagic stroke") OR ("Thromboembolism") OR ("Systemic embolism")) AND (("atrial fibrillation") OR ("non-valvular atrial fibrillation")) AND (("heart disease") OR ("myocardial infarction")) AND (("major bleeding") OR ("minor bleeding") OR ("gastrointestinal bleeding") OR ("intracranial bleeding") OR ("all-cause mortality")) AND (("comparative effectiveness") OR ("safety effectiveness")) | 29 |

**Table S2:** Quality assessment using a Modified Jadad Scoring of all the includedRCTs

| S. No | Author & Year | *Was the study described as randomized?* | *Was the method of randomization appropriate?* | *Was the study described as blinding?* | *Was the method of blinding appropriate?* | *Was there a description of withdrawals and dropouts?* | *Was there a clear description of the inclusion/e exclusion criteria?* | *Was the method used to assess adverse effects described?* | *Was the methods Of statistical Analysis described?* | *Total Score* | *Quality* |
|---|---|---|---|---|---|---|---|---|---|---|---|
| 1. | Ezekowitz et al. 2007[14] | 1 | 0 | 1 | 1 | 1 | 1 | 1 | 1 | 7 | Good |
| 2. | Connolly et al. 2009[7] | 1 | 0 | 1 | 1 | 1 | 1 | 1 | 1 | 7 | Good |
| 3. | Diener et al. 2010[15] | 1 | 0 | 1 | 0 | 0 | 1 | 1 | 1 | 5 | Fair |
| 4. | Weitz et al. 2010[16] | 1 | 0 | 1 | 0 | 0 | 1 | 1 | 1 | 5 | Fair |
| 5. | Chung et al. 2011[17] | 1 | 1 | 1 | 0 | 0 | 1 | 1 | 1 | 6 | Fair |
| 6. | Eikelboom et al. 2011 [18] | 1 | 0 | 1 | 0 | 0 | 0 | 0 | 1 | 4 | Fair |
| 7. | Fox et al. 2011 [19] | 1 | 0 | 1 | 1 | 0 | 1 | 1 | 1 | 6 | Fair |
| 8. | Granger et al. 2011[6] | 1 | 1 | 1 | 0 | 1 | 1 | 1 | 1 | 7 | Good |
| 9. | Patel et al. 2011[5] | 1 | 1 | 1 | 1 | 1 | 1 | 1 | 1 | 8 | Good |
| 10. | Easton et al. 2012[20] | 1 | 1 | 1 | 1 | 1 | 1 | 0 | 1 | 7 | Good |
| 11. | Hankey et al. 2012[21] | 1 | 1 | 1 | 1 | 1 | 1 | 1 | 1 | 7 | Good |
| 12. | Hori et al. 2012[22] | 1 | 1 | 1 | 1 | 1 | 1 | 1 | 1 | 8 | Good |
| 13. | Huisman et al. 2012[23] | 1 | 0 | 0 | 0 | 1 | 0 | 1 | 1 | 4 | Fair |
| 14. | Yamashita et al. 2012[24] | 1 | 1 | 1 | 0 | 0 | 1 | 1 | 1 | 6 | Fair |
| 15. | AI-Khatib et al. 2013[25] | 1 | 0 | 1 | 1 | 0 | 1 | 0 | 1 | 5 | Fair |
| 16. | Bahit et al. 2013[26] | 1 | 0 | 0 | 0 | 1 | 1 | 0 | 1 | 4 | Fair |
| 17. | Diepen et al. 2013[27] | 1 | 0 | 1 | 1 | 0 | 1 | 0 | 1 | 5 | Fair |
| 18. | Flaker et al. 2013[28] | 1 | 0 | 1 | 1 | 0 | 1 | 0 | 1 | 5 | Fair |
| 19. | Garcia et al. 2013[29] | 1 | 1 | 1 | 1 | 0 | 1 | 0 | 1 | 6 | Fair |
| 20. | Giugliano et al. 2013[8] | 1 | 1 | 1 | 1 | 1 | 1 | 1 | 1 | 8 | Good |
| 21. | Mahaffey et al. 2013[30] | 1 | 1 | 1 | 1 | 0 | 0 | 0 | 1 | 5 | Fair |
| 22. | Tanahashi et al. 2013 [31] | 1 | 1 | 1 | 0 | 0 | 1 | 1 | 1 | 6 | Fair |
| 23. | Alexander et al. 2014[32] | 1 | 1 | 1 | 1 | 0 | 0 | 0 | 1 | 5 | Fair |
| 24. | Breithardt et al 2014[33] | 1 | 0 | 1 | 1 | 0 | 1 | 0 | 1 | 5 | Fair |
| 25. | Cappato et al. 2014[34] | 1 | 0 | 0 | 0 | 0 | 1 | 1 | 1 | 4 | Fair |
| 26. | Halperin et al. 2014 [35] | 1 | 1 | 1 | 1 | 1 | 1 | 0 | 1 | 7 | Good |
| 27. | Mao et al. 2014[36] | 1 | 0 | 1 | 1 | 0 | 1 | 1 | 1 | 6 | Fair |
| 28. | Avezum et al. 2015[37] | 1 | 0 | 0 | 0 | 0 | 1 | 0 | 1 | 3 | Fair |
| 29. | Duraes et al. 2016[38] | 1 | 1 | 0 | 0 | 0 | 1 | 0 | 1 | 4 | Fair |
| 30. | Gibson et al. 2016[39] | 1 | 1 | 0 | 0 | 1 | 1 | 1 | 1 | 6 | Fair |
| 31. | Goette et al. 2016[40] | 1 | 1 | 1 | 1 | 1 | 1 | 1 | 1 | 8 | Good |
| 32. | Rost et al. 2016[41] | 1 | 0 | 1 | 1 | 0 | 1 | 1 | 1 | 6 | Fair |

| | | | | | | | | | | | |
|---|---|---|---|---|---|---|---|---|---|---|---|
| 33. | Lauw et al. 2017[42] | 1 | 0 | 0 | 0 | 0 | 1 | 0 | 1 | 3 | Fair |
| 34. | Ezekowitz et al. 2018 [43] | 1 | 1 | 0 | 0 | 1 | 1 | 1 | 1 | 6 | Fair |
| 35. | Kimura et al. 2018 [44] | 1 | 1 | 1 | 1 | 0 | 1 | 1 | 1 | 7 | Good |
| 36. | Kirchhof et al. 2018[45] | 1 | 0 | 1 | 1 | 1 | 1 | 1 | 1 | 7 | Good |
| 37. | Guimaraes et al. 2019[46] | 1 | 1 | 0 | 0 | 0 | 1 | 0 | 1 | 4 | Fair |
| 38. | Vranckx et al 2019[47] | 1 | 1 | 0 | 0 | 1 | 1 | 1 | 1 | 6 | Fair |
| 39. | Guimaraes et al. 2020[48] | 1 | 1 | 1 | 0 | 1 | 1 | 1 | 1 | 7 | Good |
| 40. | Mieghem et al. 2021[49] | 1 | 0 | 0 | 0 | 1 | 1 | 1 | 1 | 5 | Fair |
| 41. | Ahasan et.al. 2022[50] | 1 | 0 | 0 | 0 | 1 | 1 | 0 | 1 | 4 | Fair |
| 42. | Connolly et al. 2022[51] | 1 | 1 | 1 | 1 | 1 | 1 | 0 | 1 | 7 | Good |
| 43. | Pokorney et al. 2022[52] | 1 | 0 | 0 | 0 | 1 | 1 | 0 | 1 | 4 | Fair |
